# Interrater Reliability of Clinical Resting-State fMRI Network Classification: Comparison of Human Raters and an AI Algorithm

**DOI:** 10.64898/2026.11.24.26368595

**Authors:** Albert B. Kang, William R. Reuther, Belfin Robinson, Meitra H. Kazemi, Varina L. Boerwinkle

## Abstract

**Purpose:** Accurate classification of intrinsic brain networks is essential for the clinical interpretation of resting-state functional MRI (rs-fMRI), especially in complex pediatric populations. Manual classification of resting-state networks (RSNs) remains the clinical gold standard, yet variability among raters poses a challenge for reproducibility. This study aims to assess interrater reliability among five human raters and one AI algorithm.

**Methods:** Rs-fMRI data were collected from 269 scans in a clinical cohort predominantly composed of pediatric patients. Raters used an independent component analysis (ICA)-based pipeline to categorize RSNs into one of 13 categories. Percent agreement, Cohen’s Kappa, and Fleiss’s Kappa were used to measure agreement.

**Results:** Overall pairwise agreement for human-human pairs ranged from substantial to almost perfect (*κ* = 0.655–0.845), while human-AI agreement ranged from fair to moderate (*κ* = 0.289–0.401). Agreement was highest for Typical networks and Noise, whereas Atypical networks showed lower agreement, with moderate human-human agreement (*κ* = 0.468–0.541) and poor-to-fair human-AI agreement (*κ* = 0.147–0.305). Despite high human-human agreement on Noise components, human-AI kappa values for Noise were disproportionately low, a pattern that may reflect the high prevalence of Noise components and class imbalance in the model’s training data.

**Conclusion:** Our findings underscore the need for standardized classification frameworks and targeted training to improve consistency, particularly for atypical networks. This study supports the development of reproducible rs-fMRI workflows and the potential use of AI as a complementary screening tool within clinical neuroimaging workflows.

## Background

Resting-state functional magnetic resonance imaging (rs-fMRI) is a noninvasive neuroimaging technique that measures blood-oxygen-level-dependent (BOLD) signals to identify intrinsic brain networks [1]. Since rs-fMRI does not require patients to perform specific tasks, it is particularly advantageous for mapping functional connectivity in pediatric, cognitively impaired, or sedated patients [1]. Clinically, rs-fMRI is increasingly used in presurgical planning, epilepsy evaluation, and assessment of neurological disorders [2,3]. Identification of resting-state networks (RSNs) provides information about intrinsic functional organization that may contribute to clinical assessment and disease characterization [4].

Pediatric neuroimaging poses unique challenges compared with adult populations. One major challenge is head motion, as even small movements can produce spurious and spatially structured patterns in functional connectivity [5]. Sedation may reduce motion but can also introduce interpretive complications because sedative agents may alter both neuronal responses and the hemodynamic coupling underlying the BOLD signal [6]. For example, frontal language activation may be reduced under sedation, whereas temporal activation may remain more robust, complicating identification of clinically relevant networks [6,7]. Beyond motion and sedation, pediatric neuroimaging is further complicated by developmental changes in brain organization that may cause functional networks to appear less well defined than adult reference patterns [8].

Independent component analysis (ICA) is one of the primary computational methods used to decompose rs-fMRI data into spatially independent components corresponding to functional networks or noise sources [9]. Several approaches have also been developed to automate ICA component classification using machine learning methods [10,11]. Despite advances in automated approaches, manual visual inspection remains the clinical gold standard for component classification [12]. However, manual classification is subjective and time-consuming, requiring experts to repeatedly distinguish meaningful brain networks from noise [13]. This introduces potential variability between raters, making assessment of interrater reliability crucial for reproducible clinical interpretation [14].

Observed agreement, commonly reported as percent agreement, is one method of assessing interrater reliability for nominal data [14]. Cohen’s kappa and Fleiss’s kappa provide additional measures that account for agreement expected by chance [14–16]. Confidence intervals can also be calculated to characterize the precision of these reliability estimates [14].

While primary sensorimotor and visual networks have relatively consistent and reproducible spatial mappings that facilitate classification, atypical networks may demonstrate greater individual variability [17]. Standard ICA-based approaches can therefore have difficulty identifying and classifying components at both the group and individual level, particularly in pathological conditions [18]. Because atypical networks lack well-validated reference patterns, raters may rely more heavily on subjective expert judgment, potentially increasing interrater variability.

Multi-rater classification is one strategy for reducing uncertainty in ICA labeling [12]. In a standardized fMRI data quality-control study, moderate to substantial reliability was reported for clear include and exclude decisions, while lower agreement was observed for ambiguous or uncertain classifications [19]. Formal interrater reliability studies using kappa statistics for multicategory RSN classification in clinical populations, particularly those enriched for pediatric patients, remain limited.

Automated ICA classifiers, such as FMRIB’s ICA-based X-noiseifier, have shown promising accuracy but may have limited generalizability [20,21]. Although automated methods can achieve acceptable aggregate performance, they have not consistently matched expert visual classification across clinical settings [22]. To our knowledge, no prior study has directly quantified human-to-AI agreement using formal reliability statistics across a large multicategory RSN classification in a clinical cohort predominantly composed of pediatric patients.

This study aimed to evaluate the interrater reliability of ICA-based rs-fMRI network classification among human raters and to compare human classification with an AI-based classifier.

## Methods

### Study participants

A total of 269 clinical rs-fMRI scans obtained between May 11, 2022, and September 8, 2024, were included in this study. Table 1 summarizes patient demographic characteristics.

**Table 1.** Demographic Characteristics.

| **Characteristic** |  |
|---|---|
| Average number of components per participant (SD<sup>a</sup>) | 80 (17) |
| **Age group** |  |
| Infant, 0-1 year (%) | 35 (13%) |
| Pediatric, 1-18 years (%) | 147 (55%) |
| Adult, 18+ years (%) | 87 (32%) |
| **Sex** |  |
| Female (%) | 128 (48%) |
| Male (%) | 141 (52%) |
| **Sedation status** |  |
| No sedation | 168 (62%) |
| Pediatric sedation (%) | 88 (33%) |
| Adult sedation (%) | 10 (4%) |
| Anesthesia | 3 (1%) |

<sup>a</sup> SD, standard deviation.

### Human Raters

Five human raters participated in this study and reviewed the same set of rs-fMRI cases. Rater 1 (R1) was a pediatric neurologist with more than 14 years of experience in rs-fMRI ICA interpretation. Rater 2 was a laboratory technician with more than 1 year of rs-fMRI experience. Rater 3 was a neurosurgeon and postdoctoral researcher with 2 years of rs-fMRI ICA experience. Rater 4 was a laboratory technician with 2 years of experience, and Rater 5 was a laboratory programmer with 2 years of experience.

For human-human reliability analyses, Rater 1 served as the reference comparator and was compared with Raters 2–5. Because each rater in a pair reviewed the same cases, these comparisons were based on a common underlying set of rs-fMRI studies.

### AI Algorithm

The AI rater used in this study was a previously published multimodal deep learning framework [23]. The model combines a 3-dimensional convolutional neural network with long short-term memory networks to capture the spatial and temporal features of ICA components. It was trained on rs-fMRI data from 152 epilepsy patients and tested on 35 patients not overlapping with the scans used in this study. Network categories classified by the model included Association, Atypical Networks, Deep Grey, Frontal, Language, Modulating, Motor, Noise, Parietal, Temporal, and Vision. The model achieved an overall weighted accuracy of 70%. In this study, the algorithm served as the AI rater, and its classifications were compared with those of the five human raters to evaluate interrater reliability.

For comparison with human classifications, AI output categories were harmonized with the human classification scheme. The human categories Atypical and Atypical RSN were combined into a single Atypical category for broad categorical analyses. The AI algorithm did not assign components to the Other category; therefore, human-AI agreement for Other was not calculated and is reported as not applicable.

### Data acquisition

Rs-fMRI scans were acquired on a 3-Tesla MRI scanner using a 2-dimensional gradient echo-planar imaging sequence optimized for BOLD contrast. Each acquired volume comprised 64 × 64 × 32 voxels at an isotropic resolution of 4.0 × 4.0 × 4.0 mm. Scanning parameters included a repetition time of 2.01 s and an echo time of 35 ms. An interleaved slice-ordering approach was used to reduce slice-timing artifacts. Each scan consisted of 600 volumes, corresponding to a total acquisition time of approximately 20 minutes.

### Data preprocessing

Before ICA, rs-fMRI data were preprocessed using FSL FEAT version 3.13 [24]. To ensure signal equilibration, the first 5 volumes were discarded. Motion correction was performed using MCFLIRT, and slice-timing correction was applied using the interleaved acquisition order. Brain extraction was performed using BET. For spatial smoothing, a 1-mm full-width at half-maximum (FWHM) Gaussian kernel was applied to improve the signal-to-noise ratio; a relatively small kernel was selected to limit blurring into adjacent arteries and ventricles. Intensity normalization was not applied. Low-frequency drifts were removed using a 100-s high-pass temporal filter. Functional images were aligned to each patient’s T1-weighted structural MRI using FLIRT (12 degrees of freedom, 90° search space) and resampled to a 4-mm standard-space resolution. Finally, the time series was variance-normalized, noise was investigated using multi-session temporal concatenation ICA, and statistical thresholding was applied using cluster-based correction (Z = 2.3, p < 0.05).

### Rs-fMRI classification

ICA of rs-fMRI data was performed using Multivariate Exploratory Linear Optimized Decomposition into Independent Components (MELODIC). MELODIC is a probabilistic ICA model that uses Laplace approximation to determine the number of independent components, thereby eliminating the need to pre-specify the component count [25]. It generates spatial maps and corresponding time courses that capture a broad range of brain activity.

These components were then reviewed by raters to distinguish meaningful neural signal from noise, including motion-related and physiologic artifacts. Components judged to represent meaningful signal were manually categorized into the following network classes: Atypical, Atypical RSN, Motor, Language, Parietal, Frontal, Temporal, Vision, Deep Grey, Modulating, Association, Noise, and Other. Classification was based on evaluation of each component’s 3-dimensional spatial map, time-domain signal, and frequency-domain signal. Representative ICA components illustrating the major classification categories are shown in Fig. 1.

**Fig 1.**
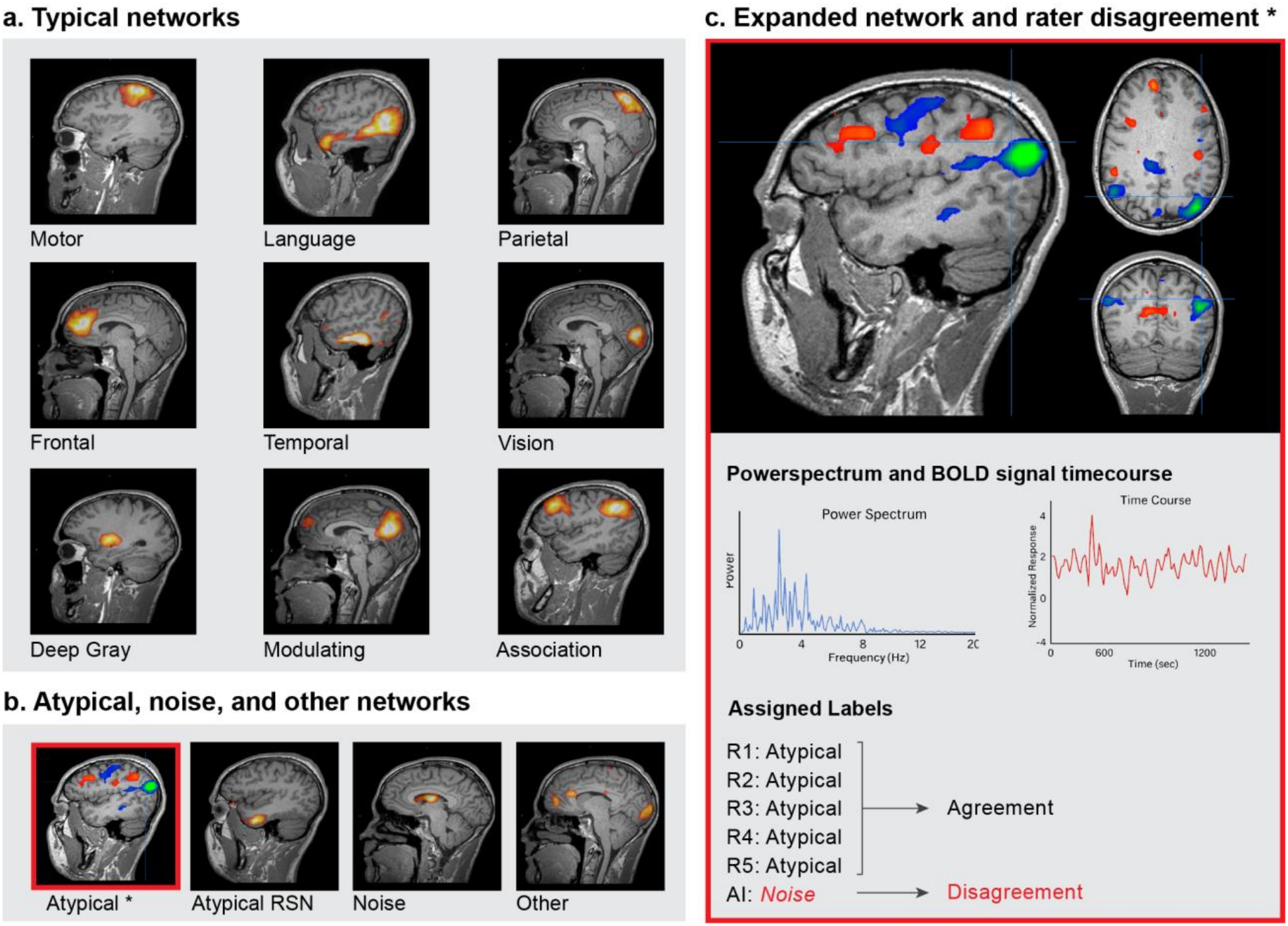
Examples of ICA component classification. Examples of rs-fMRI independent component classification. a) Example spatial maps for the nine typical resting-state network (RSN) classes. b) Example spatial maps for the four remaining classes (Atypical, Atypical RSN, Noise, and Other). c) Expanded view of the component marked with an asterisk in b, showing its spatial map, time course, power spectrum, and the differing labels assigned during human and AI review.

### Data management and statistical analysis

Data analysis was performed using R, ver. 4.5.1 (R Foundation for Statistical Computing, Vienna, Austria) within RStudio: Integrated Development Environment for R, ver. 2026.1.0.392 (Posit Software, PBC, Boston, MA, USA).

Interrater reliability was assessed using percent agreement, Cohen’s kappa, and Fleiss’s kappa. Percent agreement was calculated as the number of agreement scores divided by the total number of scores. For overall agreement, the total number of scores was equal to the total number of components. For categorical agreement, the total number of scores was defined as the number of components for which at least one rater assigned the category of interest. Cohen’s kappa was used to account for agreement expected by chance, and Fleiss’s kappa was used to assess agreement among more than 2 raters.

Pairwise agreement was assessed across 9 prespecified rater pairs: Rater 1 (R1) versus R2, R1 versus R3, R1 versus R4, R1 versus R5, R1 versus AI, R2 versus AI, R3 versus AI, R4 versus AI, and R5 versus AI. Categorical agreement was first assessed within 4 broad classifications: Typical, Atypical, Noise, and Other. Typical networks included the 9 specific classes Motor, Language, Parietal, Frontal, Temporal, Vision, Deep Grey, Modulating, and Association. Atypical included the Atypical and Atypical RSN classes. Agreement was then assessed across all 13 network classes. Agreement by sedation status was also evaluated within each sedation group. Cohen’s kappa values were interpreted using standard benchmarks as follows: poor (< 0.2), fair (0.2–0.4), moderate (0.4–0.6), substantial (0.6–0.8), and almost perfect (0.8–1.0) [15].

## Results

Table 2 summarizes overall and categorical agreement between human-human and human-AI rater pairs. Overall, all human-human pairs demonstrated at least substantial agreement, with Rater 1 and Rater 4 achieving almost perfect agreement (*κ* = 0.845). Human-AI agreement was lower, with Cohen’s kappa values ranging from 0.289 to 0.401. Rater 1 demonstrated the highest overall agreement with the AI algorithm (*κ* = 0.401), whereas Rater 3 demonstrated the lowest (*κ* = 0.289). Overall agreement across human-human and human-AI comparisons is summarized visually in Fig. 2.

**Table 2.**
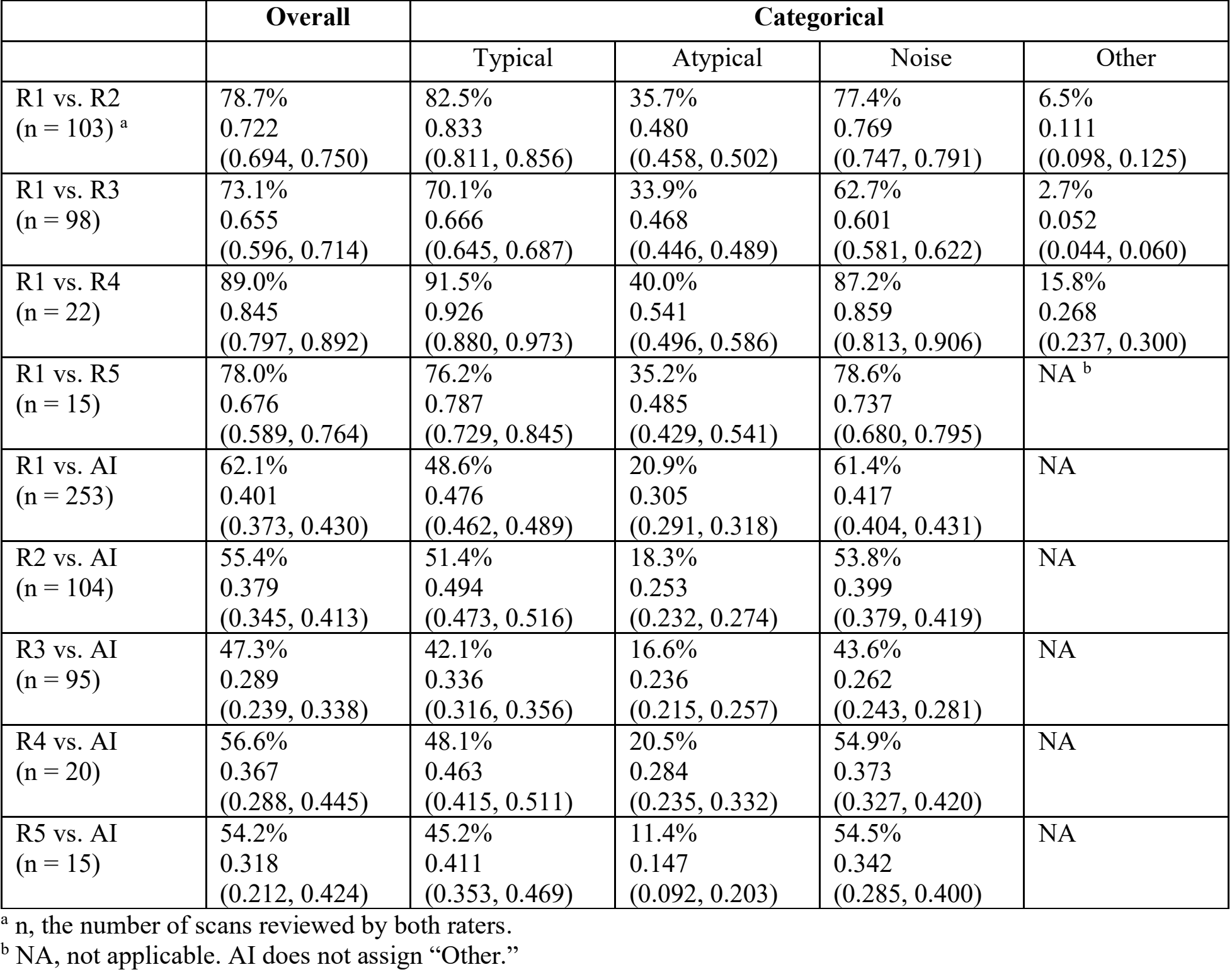
Pairwise Rater Agreement; %-agreement, Cohen’s Kappa (95% CI)

**Fig 2.**
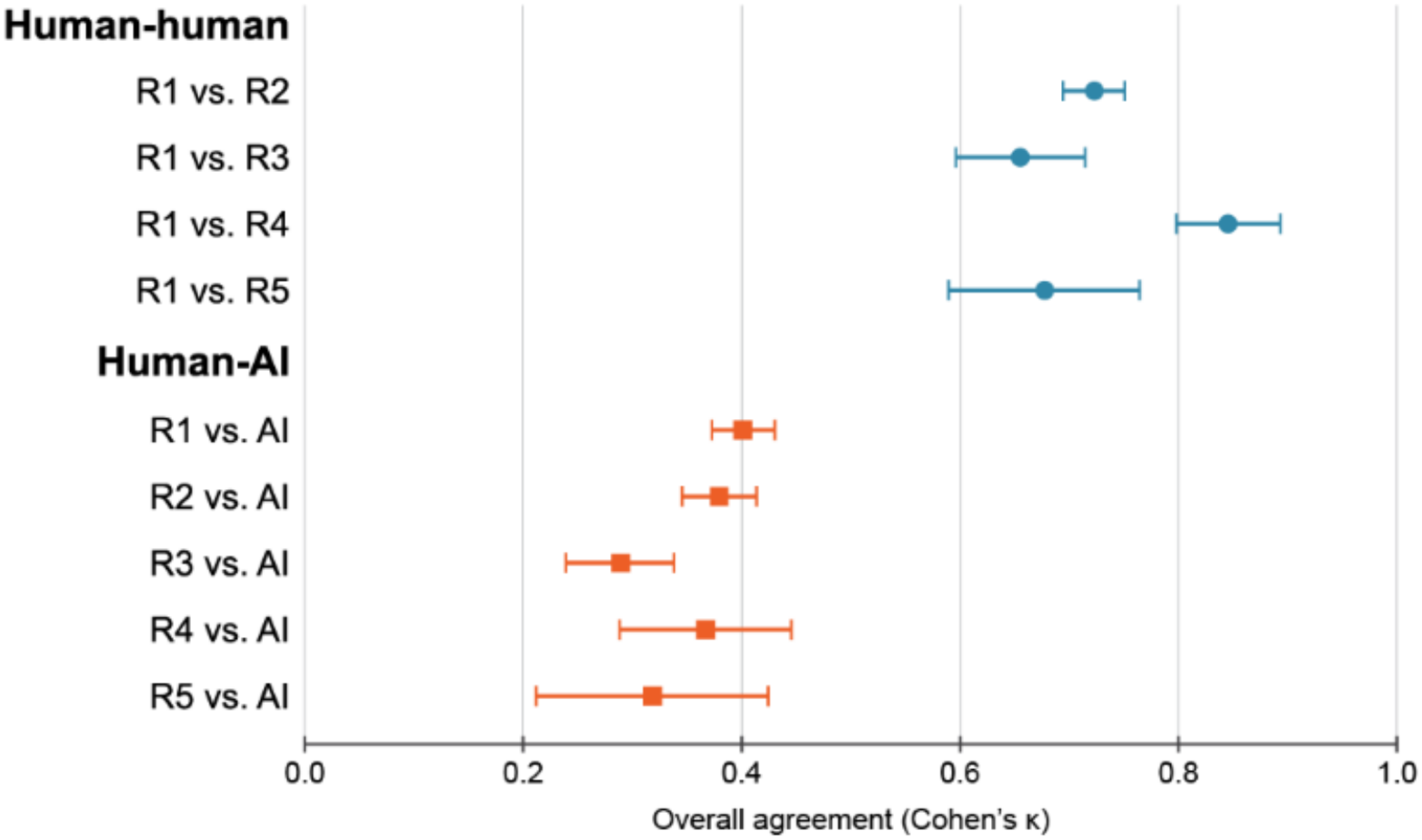
Overall human-human and human-AI pairwise agreement. Overall pairwise agreement in the classification of rs-fMRI independent components. Circles indicate comparisons between human raters, and squares indicate comparisons between each human rater and the AI classifier. Each point represents Cohen’s *κ*, with horizontal bars showing the 95% confidence interval.

Categorical agreement was assessed across four broad categories: Typical, Atypical, Noise, and Other, with results for all 13 individual classes provided in Online Resource 1. Among human-human pairs, agreement was highest for Typical and Noise components. R1 versus R2 demonstrated almost perfect agreement for Typical networks (*κ* = 0.833), while R1 versus R4 demonstrated almost perfect agreement for both Typical networks (*κ* = 0.926) and Noise (*κ* = 0.859). Agreement for Atypical networks was consistently lower across human-human comparisons, with kappa values ranging from 0.468 to 0.541.

A similar pattern was observed for human-AI comparisons. Typical networks generally demonstrated the highest human-AI kappa values, ranging from 0.336 to 0.494, whereas Atypical networks demonstrated lower agreement, with kappa values ranging from 0.147 to 0.305. Although percent agreement for Noise components was relatively high across human-AI comparisons, the corresponding kappa values were lower than those observed for Typical networks. This discrepancy between percent agreement and kappa was observed consistently across human-AI rater pairs.

Multi-rater agreement among two human raters and the AI algorithm is summarized in Table 3. Across all four three-rater combinations, Fleiss’s kappa values were within the moderate agreement range, ranging from 0.438 to 0.555. The R1 versus R4 versus AI combination demonstrated the highest agreement (*κ* = 0.555), whereas R1 versus R3 versus AI demonstrated the lowest agreement (*κ* = 0.438).

**Table 3.** Multi-rater Agreement.

|  | R1 vs. R2 vs. AI<br>(n = 100)<sup>a</sup> | R1 vs. R3 vs. AI<br>(n = 94) | R1 vs. R4 vs. AI<br>(n = 19) | R1 vs. R5 vs. AI<br>(n = 15) |
|---|---|---|---|---|
| %-agreement | 52.7% | 43.5% | 55.9% | 50.1% |
| Fleiss's Kappa<br>(95% CI) | 0.522<br>(0.495, 0.548) | 0.438<br>(0.392, 0.485) | 0.555<br>(0.508, 0.603) | 0.456<br>(0.365, 0.548) |

<sup>a</sup> n, the number of scans reviewed by all three raters.

Agreement by sedation status is presented in Table 4. For R1 versus R2, agreement was similar between the no-sedation group (*κ* = 0.718) and the pediatric sedation group (*κ* = 0.729). For R1 versus R3, agreement was *κ* = 0.630 in the no-sedation group and *κ* = 0.719 in the pediatric sedation group. Lower agreement was observed for R1 versus R3 in the adult sedation group (*κ* = 0.332). However, the adult sedation and anesthesia subgroups contained very small numbers of patients and were therefore interpreted descriptively.

**Table 4.** Agreement by Sedation Status; %-agreement, Cohen’s Kappa (95% CI)

| Sedation Status | R1 vs. R2 (n = 101) | R1 vs. R3 (n = 88) |
|---|---|---|
| No sedation (n = 63, 54)<sup>a</sup> | 79.0%<br>0.718<br>(0.707, 0.730) | 71.6%<br>0.630<br>(0.619, 0.641) |
| Pediatric sedation (n = 36, 30) | 78.1%<br>0.729<br>(0.715, 0.743) | 77.1%<br>0.719<br>(0.704, 0.734) |
| Adult sedation (n = 2, 3) | 77.3%<br>0.542<br>(0.443, 0.642) | 54.5%<br>0.332<br>(0.281, 0.383) |
| Anesthesia (n = 0, 1) | NA | 89.6%<br>0.852<br>(0.744, 0.961) |

<sup>a</sup> n, the number of scans reviewed by both raters per sedation group for R1 vs. R2 and R1 vs. R3, respectively.

## Discussion

This study evaluated interrater reliability among five human raters and one AI algorithm for ICA-based rs-fMRI network classification in a clinical cohort predominantly composed of pediatric patients. Human-human pairwise agreement ranged from substantial to almost perfect, indicating that trained raters can achieve high levels of consistency when using a structured classification framework. In contrast, human-AI agreement ranged from fair to moderate, and the addition of the AI algorithm to human rater pairs reduced multi-rater agreement into the moderate range. These findings highlight a persistent gap between expert human classification and automated classification in clinical rs-fMRI.

Categorical analyses demonstrated that agreement was strongest for Typical networks and Noise, whereas Atypical networks were more difficult to classify for both human raters and the AI algorithm. Human-human kappa values for Atypical networks were consistently lower than those observed for Typical networks and Noise. This finding is consistent with the greater spatial variability and reduced availability of stable reference patterns for atypical network organization, particularly in patients with neurological pathology [12]. Greater standardization of criteria for atypical network classification may therefore improve reproducibility across raters.

A notable discrepancy was observed between percent agreement and Cohen’s kappa for Noise in human-AI comparisons. Although percent agreement for Noise was relatively high, corresponding kappa values were lower than those observed for Typical networks. This pattern is consistent with a prevalence effect and may be influenced by the high proportion of Noise components in the AI model’s training dataset [23]. Because Noise represented approximately half of the ICA components in the training data, class imbalance may have influenced the model’s classification behavior. These findings suggest that automated identification of Noise should be interpreted cautiously in clinical workflows, particularly when misclassification could result in clinically relevant components being excluded from further review.

Sedation status did not appear to substantially alter agreement between the two rater pairs evaluated in the larger no-sedation and pediatric-sedation groups. Agreement remained substantial for R1 versus R2 in both groups and was also substantial for R1 versus R3 in the pediatric-sedation group. Findings from the adult-sedation and anesthesia subgroups should be considered exploratory because of the very small number of patients represented in these groups. Larger cohorts would be required to determine whether sedation meaningfully influences rs-fMRI network classification reliability.

These findings support a complementary role for AI in clinical rs-fMRI workflows rather than use as a standalone classifier. Human raters achieved agreement levels consistent with prior studies of ICA-based classification [20], and multi-rater consensus approaches have been recommended for managing component-level ambiguity [12]. The fair-to-moderate agreement observed between individual human raters and the AI algorithm suggests that automated classification may be most useful as a screening or first-pass tool that identifies components for subsequent expert review, particularly for well-defined network categories.

Several limitations should be considered. This was a single-center retrospective study using a clinical cohort with a pediatric majority but a substantial adult component. Human raters differed in professional background and experience, and not all possible human-human rater pairings were evaluated. In addition, each patient contributed multiple ICA components, creating clustered observations within individual patients. The adult-sedation and anesthesia subgroups were very small, limiting interpretation of those analyses. Finally, the AI algorithm was developed using data from the same institution, which may limit generalizability to other populations, scanners, or preprocessing pipelines.

In conclusion, trained human raters demonstrated substantial to almost perfect agreement when classifying ICA-based rs-fMRI components using a structured framework, whereas agreement between human raters and the AI algorithm was lower. Atypical networks remained the most challenging category for both human and automated classification. These findings support continued use of expert human review as the clinical reference standard while suggesting a potential complementary role for AI in assisting with well-defined network classifications. Further standardization of atypical network criteria and external validation of automated classifiers will be important for improving the reliability and generalizability of AI-assisted rs-fMRI interpretation.

## Supporting information

Online Resource 1

## Data Availability

The data and scripts that support the findings of this study are not publicly available due to patient privacy restrictions and institutional data use agreements, but are available from the corresponding author upon reasonable request and with appropriate institutional approval.

## Statements and Declarations

### Ethics approval and consent to participate

This study followed the ethical standards of the 1964 Declaration of Helsinki and its later amendments. The Institutional Review Board of the University of North Carolina at Chapel Hill (IRB #24-0833) approved this study. All rs-fMRI datasets were obtained under informed consent from participants, and all methods were carried out in accordance with relevant guidelines and regulations.

## Author Contributions

**Albert Kang:** Writing – original draft, Data curation, Investigation, Formal analysis. **William Reuther:** Writing – review and editing, Data curation, Project administration. **Belfin Robinson:** Writing – review and editing, Methodology, Software. **Meitra Kazemi:** Writing – review and editing, Project administration. **Varina Boerwinkle:** Writing – review and editing, Conceptualization, Data curation, Supervision.

## Acknowledgements

We sincerely thank the patients that participated in this study. We also acknowledge the research assistants for their assistance with clinical data acquisition.

