## Supplementary material for "Interrater Reliability of Clinical Resting-State fMRI Network Classification: Comparison of Human Raters and an AI Algorithm": Online Resource 1

Journal: Clinical Neuroradiology

**Online Resource 1** Pairwise rater agreement, overall and categorical across all 13 RSN categories. %-agreement, Cohen's Kappa, (95% Confidence Interval)

| | R1 vs. R2<br>(n = 103) [a] | R1 vs. R3<br>(n = 98) | R1 vs. R4<br>(n = 22) | R1 vs. R5<br>(n = 15) | R1 vs. AI<br>(n = 253) | R2 vs. AI<br>(n = 104) | R3 vs. AI<br>(n = 95) | R4 vs. AI<br>(n = 20) | R5 vs. AI<br>(n = 15) |
|---|---|---|---|---|---|---|---|---|---|
| Overall | 78.7%<br>0.722<br>(0.694, 0.750) | 73.1%<br>0.655<br>(0.596, 0.714) | 89.0%<br>0.845<br>(0.797, 0.892) | 78.0%<br>0.676<br>(0.589, 0.764) | 62.1%<br>0.401<br>(0.373, 0.430) | 55.4%<br>0.379<br>(0.345, 0.413) | 47.3%<br>0.289<br>(0.239, 0.338) | 56.6%<br>0.367<br>(0.288, 0.445) | 54.2%<br>0.318<br>(0.212, 0.424) |
| Categorical | | | | | | | | | |
| Atypical | 31.5%<br>0.439<br>(0.418, 0.461) | 33.4%<br>0.463<br>(0.442, 0.484) | 32.4%<br>0.463<br>(0.419, 0.508) | 23.3%<br>0.342<br>(0.289, 0.395) | 21.4%<br>0.314<br>(0.301, 0.328) | 18.8%<br>0.264<br>(0.243, 0.286) | 16.4%<br>0.233<br>(0.212, 0.254) | 18.0%<br>0.252<br>(0.203, 0.302) | 12.1%<br>0.160<br>(0.104, 0.216) |
| Atypical RSN | 10.5%<br>0.177<br>(0.155, 0.199) | 9.1%<br>0.166<br>(0.147, 0.185) | 42.2%<br>0.586<br>(0.541, 0.632) | 4.0%<br>0.068<br>(0.014, 0.122) | 1.1%<br>0.021<br>(0.016, 0.025) | 0%<br>-0.001<br>(-0.007, 0.006) | 0%<br>0.000<br>(-0.019, 0.019) | 0%<br>-0.001<br>(-0.019, 0.016) | NA |
| Motor | 75.9%<br>0.854<br>(0.832, 0.876) | 62.4%<br>0.755<br>(0.733, 0.776) | 86.0%<br>0.921<br>(0.875, 0.967) | 53.5%<br>0.682<br>(0.624, 0.740) | 39.4%<br>0.543<br>(0.529, 0.556) | 41.9%<br>0.566<br>(0.544, 0.588) | 30.4%<br>0.438<br>(0.416, 0.459) | 42.3%<br>0.575<br>(0.525, 0.624) | 31.9%<br>0.456<br>(0.399, 0.512) |
| Language | 71.9%<br>0.821<br>(0.799, 0.843) | 62.0%<br>0.751<br>(0.729, 0.773) | 75.0%<br>0.848<br>(0.801, 0.894) | 62.1%<br>0.755<br>(0.697, 0.813) | 28.9%<br>0.419<br>(0.406, 0.433) | 26.4%<br>0.380<br>(0.360, 0.401) | 26.0%<br>0.382<br>(0.360, 0.404) | 21.4%<br>0.319<br>(0.271, 0.368) | 26.4%<br>0.393<br>(0.335, 0.451) |
| Parietal | 69.8%<br>0.814<br>(0.792, 0.836) | 65.7%<br>0.780<br>(0.759, 0.802) | 86.4%<br>0.924<br>(0.877, 0.970) | 56.5%<br>0.713<br>(0.655, 0.771) | 22.0%<br>0.333<br>(0.319, 0.347) | 21.2%<br>0.321<br>(0.299, 0.343) | 21.3%<br>0.318<br>(0.297, 0.340) | 13.5%<br>0.218<br>(0.174, 0.261) | 16.1%<br>0.254<br>(0.196, 0.312) |
| Frontal | 64.4%<br>0.771<br>(0.749, 0.794) | 61.8%<br>0.742<br>(0.720, 0.764) | 86.9%<br>0.925<br>(0.878, 0.971) | 59.0%<br>0.727<br>(0.668, 0.785) | 19.3%<br>0.300<br>(0.288, 0.312) | 21.7%<br>0.333<br>(0.312, 0.354) | 12.9%<br>0.203<br>(0.187, 0.218) | 32.4%<br>0.458<br>(0.410, 0.506) | 10.1%<br>0.157<br>(0.105, 0.210) |
| Temporal | 51.3%<br>0.668<br>(0.646, 0.690) | 53.5%<br>0.682<br>(0.660, 0.703) | 85.1%<br>0.916<br>(0.870, 0.963) | 45.3%<br>0.610<br>(0.553, 0.668) | 12.0%<br>0.192<br>(0.178, 0.206) | 12.4%<br>0.200<br>(0.179, 0.222) | 9.0%<br>0.130<br>(0.109, 0.151) | 16.5%<br>0.257<br>(0.208, 0.306) | 9.5%<br>0.139<br>(0.081, 0.197) |
| Vision | 83.1%<br>0.902<br>(0.879, 0.924) | 73.7%<br>0.839<br>(0.817, 0.860) | 92.8%<br>0.960<br>(0.914, 1.000) | 73.5%<br>0.839<br>(0.781, 0.897) | 38.6%<br>0.530<br>(0.516, 0.543) | 45.8%<br>0.604<br>(0.582, 0.626) | 32.3%<br>0.454<br>(0.432, 0.476) | 43.6%<br>0.580<br>(0.531, 0.630) | 37.4%<br>0.513<br>(0.456, 0.571) |
| Deep Grey | 70.5% | 58.6% | 87.7% | 56.5% | 24.6% | 27.5% | 18.4% | 23.8% | 14.0% |

| | | | | | | | | | |
|---|---|---|---|---|---|---|---|---|---|
| | 0.819<br>(0.797, 0.841) | 0.727<br>(0.705, 0.748) | 0.931<br>(0.885, 0.978) | 0.705<br>(0.648, 0.762) | 0.375<br>(0.361, 0.388) | 0.411<br>(0.390, 0.433) | 0.285<br>(0.264, 0.306) | 0.365<br>(0.318, 0.413) | 0.211<br>(0.157, 0.265) |
| Modulating | 67.5%<br>0.801<br>(0.779, 0.823) | 75.3%<br>0.856<br>(0.834, 0.878) | 87.5%<br>0.932<br>(0.885, 0.978) | 70.0%<br>0.819<br>(0.761, 0.878) | 32.6%<br>0.483<br>(0.469, 0.496) | 32.7%<br>0.482<br>(0.461, 0.504) | 34.0%<br>0.499<br>(0.477, 0.521) | 21.2%<br>0.336<br>(0.288, 0.385) | 43.3%<br>0.597<br>(0.539, 0.655) |
| Association | 57.6%<br>0.724<br>(0.702, 0.746) | 26.8%<br>0.402<br>(0.383, 0.420) | 78.8%<br>0.879<br>(0.833, 0.926) | 55.0%<br>0.706<br>(0.648, 0.763) | 28.3%<br>0.429<br>(0.415, 0.443) | 21.0%<br>0.331<br>(0.309, 0.352) | 15.3%<br>0.244<br>(0.226, 0.261) | 16.7%<br>0.275<br>(0.225, 0.324) | 26.5%<br>0.408<br>(0.350, 0.465) |
| Other | 6.5%<br>0.111<br>(0.098, 0.125) | 2.7%<br>0.052<br>(0.044, 0.060) | 15.8%<br>0.268<br>(0.237, 0.300) | NA [b] | NA | NA | NA | NA | NA |
| Noise | 77.4%<br>0.769<br>(0.747, 0.791) | 62.7%<br>0.601<br>(0.581, 0.622) | 87.2%<br>0.859<br>(0.813, 0.906) | 78.6%<br>0.737<br>(0.680, 0.795) | 61.4%<br>0.417<br>(0.404, 0.431) | 53.8%<br>0.399<br>(0.379, 0.419) | 43.6%<br>0.262<br>(0.243, 0.281) | 54.9%<br>0.373<br>(0.327, 0.420) | 54.5%<br>0.342<br>(0.285, 0.400) |

[a] n, the number of scans reviewed by both raters.

[b] NA, not applicable. Rater 5 and AI Algorithm did not assign "Other."
